# Greater risk of severe COVID-19 in non-White ethnicities is not explained by cardiometabolic, socioeconomic, or behavioural factors, or by 25(OH)-vitamin D status: study of 1,326 cases from the UK Biobank

**DOI:** 10.1101/2020.06.01.20118943

**Authors:** Zahra Raisi-Estabragh, Celeste McCracken, Mae S. Bethell, Jackie Cooper, Cyrus Cooper, Mark J. Caulfield, Patricia B. Munroe, Nicholas C. Harvey, Steffen E. Petersen

## Abstract

**Background:** We examined whether the greater severity of coronavirus disease 2019 (COVID-19) amongst men and non-White ethnicities is explained by cardiometabolic, socio-economic, or behavioural factors.

**Methods:** We studied 4,510 UK Biobank participants tested for COVID-19 (positive, *n* = 1,326). Multivariate logistic regression models including age, sex, and ethnicity were used to test whether addition of: 1)cardiometabolic factors (diabetes, hypertension, high cholesterol, prior myocardial infarction, smoking, BMI); 2)25(OH)-vitamin D; 3)poor diet; 4)Townsend deprivation score; 5)housing (home type, overcrowding); or 6)behavioural factors (sociability, risk taking) attenuated sex/ethnicity associations with COVID-19 status.

**Results:** There was over-representation of men and non-White ethnicities in the COVID-19 positive group. Non-Whites had, on average, poorer cardiometabolic profile, lower 25(OH)-vitamin D, greater material deprivation, and were more likely to live in larger households and flats/apartments. Male sex, non-White ethnicity, higher BMI, Townsend deprivation score, and household overcrowding were independently associated with significantly greater odds of COVID-19. The pattern of association was consistent for men and women; cardiometabolic, socio-demographic and behavioural factors did not attenuate sex/ethnicity associations.

**Conclusions:** Sex and ethnicity differential pattern of COVID-19 is not adequately explained by variations in cardiometabolic factors, 25(OH)-vitamin D levels, or socio-economic factors. Investigation of alternative biological pathways and different genetic susceptibilities is warranted.

## Introduction

The coronavirus disease 2019 (COVID-19) pandemic has to date resulted in over 5.8 million cases and 362,000 deaths worldwide^1^. An increasing number of reports have highlighted men and Black and Minority Ethnic (BAME) cohorts as at higher risk of adverse COVID-19 outcomes^2,3^. Variations in cardiometabolic disease burden^4^, oestrogen pathway activity^5^, vitamin D levels^6^, and angiotensin converting enzyme (ACE) 2 receptor expression^7^ have been proposed as potential explanations for the differential pattern of disease severity. Furthermore, disparities in socioeconomic standards, housing conditions, socialisation habits, and risk perception are potentially relevant with likely implications for risk of exposure and transmission. Understanding the significance of these factors is urgently needed to inform public health and research efforts.

We therefore investigated, in the UK Biobank (UKB) cohort, whether differential patterns of COVID-19 incidence and severity by sex and ethnicity might be explained by cardiometabolic, socioeconomic, lifestyle, and behavioural exposures.

## Methods

### Setting and study population

UKB is a prospective cohort study of over half a million men and women from across the UK covering a range of urban and rural settings. Recruitment was between 2006–2010 through postal invite of individuals aged 40–69 years-old identified through National Health Service (NHS) registers. Baseline assessment included detailed characterisation of socio-demographics, lifestyle, health, a series of physical measures, and blood biochemistry. The protocol is publicly available^8^. Data linkage with Hospital Episode Statistics (HES) enables prospective tracking of health outcomes for all participants with conditions recorded according to international classification of disease (ICD). Incidence of key events, such as myocardial infarction (MI), are algorithmically defined by crosschecking over multiple data sources^9^. Linkage with Public Health England infection data has enabled rapid release of linked COVID-19 test results for UKB participants to researchers^10^. As such, near-real time study of the biological and epidemiological determinants of the disease is possible. The latest data release (29/05/2020) includes test results from 16/03/2020 to 18/05/2020. As UK testing during this period was almost entirely restricted to hospitalised patients, researchers have been advised that COVID-19 positive status in UKB can be taken as surrogate for severe disease^11^.

### Exposures

We considered relevant demographic (age, sex, ethnicity), biological (cardiometabolic, 25(OH)-vitamin D status, poor diet quality), socio-economic (material deprivation, type of home, household overcrowding), and behavioural (sociability, attitude to risk) disease determinants (Supplementary Table 1).

We used age and sex as recorded at baseline. Ethnicity is reported as Caucasian (any White background) and non-Caucasian. For consistency with wider UK classification, we document ethnicity as White and non-White. For the latter we report breakdown of ethnicities as per existing UKB categories: Black (Caribbean, African, any other Black background), Asian (Indian, Pakistani, Bangladeshi, any other Asian background), Chinese, Mixed (White and Black Caribbean, White and Black African, White and Asian, Any other mixed background), and “other”. Townsend deprivation score is reported by the UKB as a measure of material deprivation calculated at baseline: zero, positive, and negative scores correspond to average, higher and lower levels of deprivation respectively, relative to national averages^12^. We used type of housing as a binary variable comprising communal living spaces (flat, apartment, sheltered accommodation) vs stand-alone housing (house, bungalow). We considered household overcrowding based on self-report of household size (number of people in the same home) and intergenerational cohabitation (number of generations in the same home). Socialisation habits were defined per self-reports of frequency of family/friend visits and participation in regular leisure activities outside the home (sports, pub, religious group, adult education class). Attitude to risk was assessed using self-report of tendency “to take risks” (binary yes/no). BMI was calculated from height and weight recorded at baseline. Smoking status was based on self-report. Hypertension, diabetes, and hypercholesterolaemia were defined through cross-checking across self-report and HES data. A list of ICD codes used is available in Supplementary Table 2. Prior MI was obtained from UKB algorithmically defined health outcomes. We used serum 25(OH)-vitamin D levels measured at baseline (CLIA analysis on a DiaSorin Ltd. LIASON XL), limiting to results between 10-375 nmol/L based on the manufacturer’s analytic reportable range^13^. We adjusted for seasonality by regressing vitamin D on month of sampling as a factor, this allowed derivation of vitamin D adjusted to the same month for each participant. Preliminary analysis demonstrated significant difference in vitamin D by ethnicity and variation in the degree of seasonal variation by ethnicity (Figure 1D). We therefore performed seasonality adjustment separately for White and non-White ethnicities and added the intercept to the adjusted variables to maintain the difference between the two ethnic groups (i.e. regression residuals + intercept). We used processed meat intake as a marker of poor diet quality. We converted self-reported weekly intake frequencies into probabilities of daily intake and multiplied by portion size to derive a continuous measure of daily consumption in grams, as per previous publications using this dataset^14,15^.

**Figure 1:**
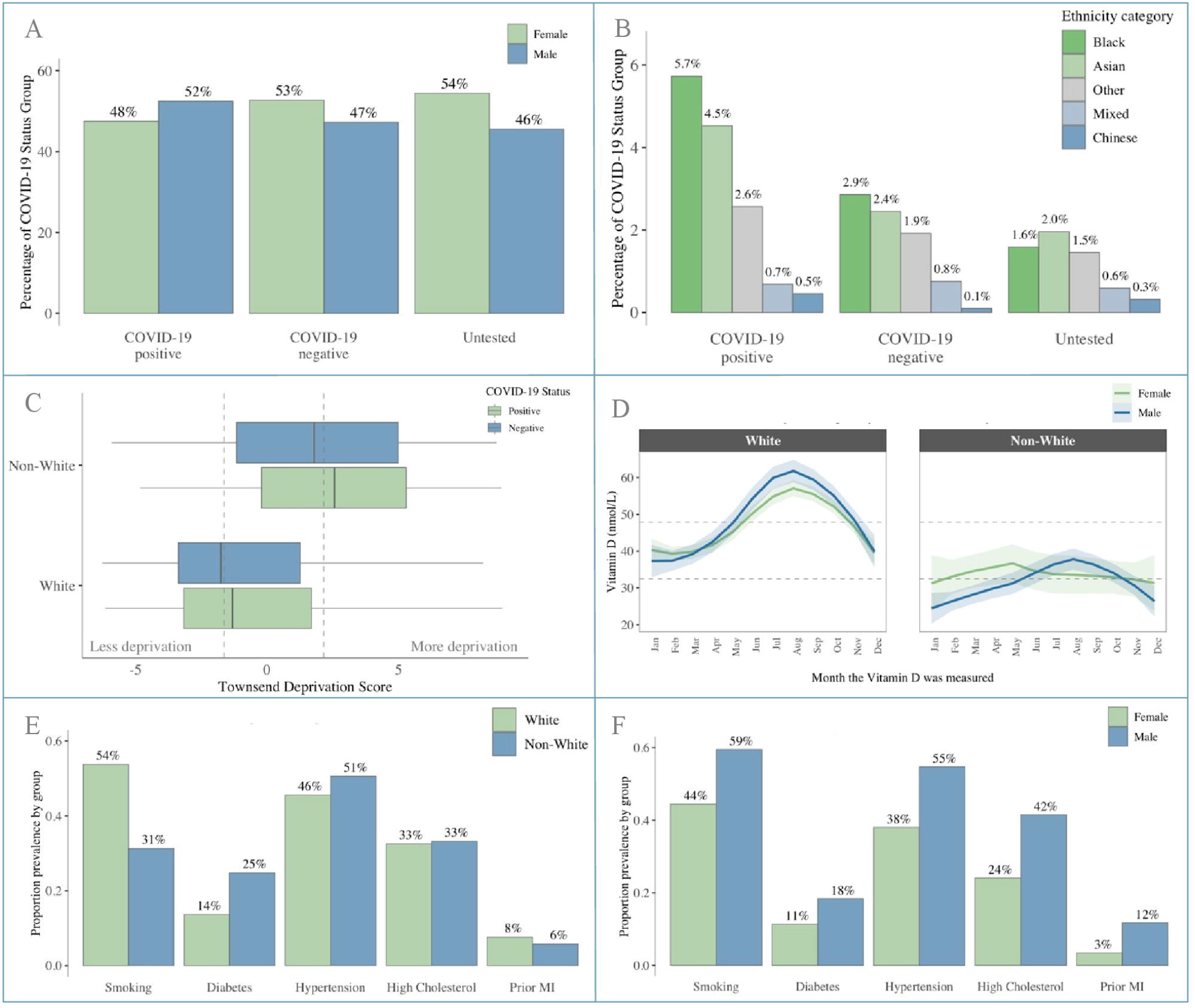
Baseline participant characteristics.

**Figure 1 footnote: Panel A:** Male: Female split by COVID-19 status; **Panel B:** Percentage of participants from different non-Caucasian ethnicities by COVID-19 status; **Panel C:** Townsend deprivation score by ethnicity and COVID-19 status; **Panel D:** Vitamin D levels by month of measurement stratified by sex and ethnicity; **Panel E:** Cardiometabolic profile stratified by ethnicity; **Panel F:** Cardiometabolic profile stratified by sex. COVID-19: coronavirus disease 2019; MI: myocardial infarction.

### Ethics

This study was covered by the ethics approval for UKB studies from the NHS National Research Ethics Service on 17th June 2011 (Ref 11/NW/0382) and extended on 10th May 2016 (Ref

16/NW/0274).

### Statistical analysis

Statistical analysis was performed using R Version 3.6.2 [R Core Team (2019). R: A language and environment for statistical computing. R Foundation for Statistical Computing, Vienna, Austria. URL https://www.R-project.org/], and RStudio Version 1.2.5019 [RStudio Team (2015). RStudio: Integrated Development for R. RStudio, Inc., Boston, MA URL http://www.rstudio.com/].

UKB participants were grouped according to COVID-19 status: test positive, test negative, and untested. In analysis of an earlier data release, we demonstrated similar associations when comparing the untested cohort with both the test negatives and test positives; suggesting that comparison with the whole cohort in this way reveals associations with general hospitalisation rather than specifically with COVID-19^16^. Therefore, to avoid bias relating to hospitalisation, in the present study, we limited logistic regression models to within the tested cohort i.e. test positives vs test negatives. We performed analyses in the whole tested sample, and separately in men and women. Logistic regression models were used to examine univariate and multivariate associations. We undertook individual multivariate models for each hypothesis to maximise sample size. We tested for multicollinearity setting a variance inflation factor (VIF) cut-off of 2.5. We present odds ratio (OR) for each exposure with the corresponding 95% confidence interval (CI) and p-value.

## RESULTS

### Population characteristics

#### Sex and ethnicity

Test results for 4,510 participants were available (positive, *n* = 1,326; negative, *n* = 3,184). Baseline characteristics are summarised in Table 1. Comparisons with the untested cohort (*n* = 497,996) and characteristics by sex and ethnicity are summarised in Supplementary Tables 3, 4 and 5. There was over-representation of men and non-White ethnicities in the test positive cohort (Figure 1A, Figure 1B). Individuals of Black and Asian ethnicity were most disproportionately affected with Black ethnicities contributing over 3.5 times the number of positive cases than their representation in the untested cohort (Supplementary Table 3, Figure 1B).

**Table 1.**
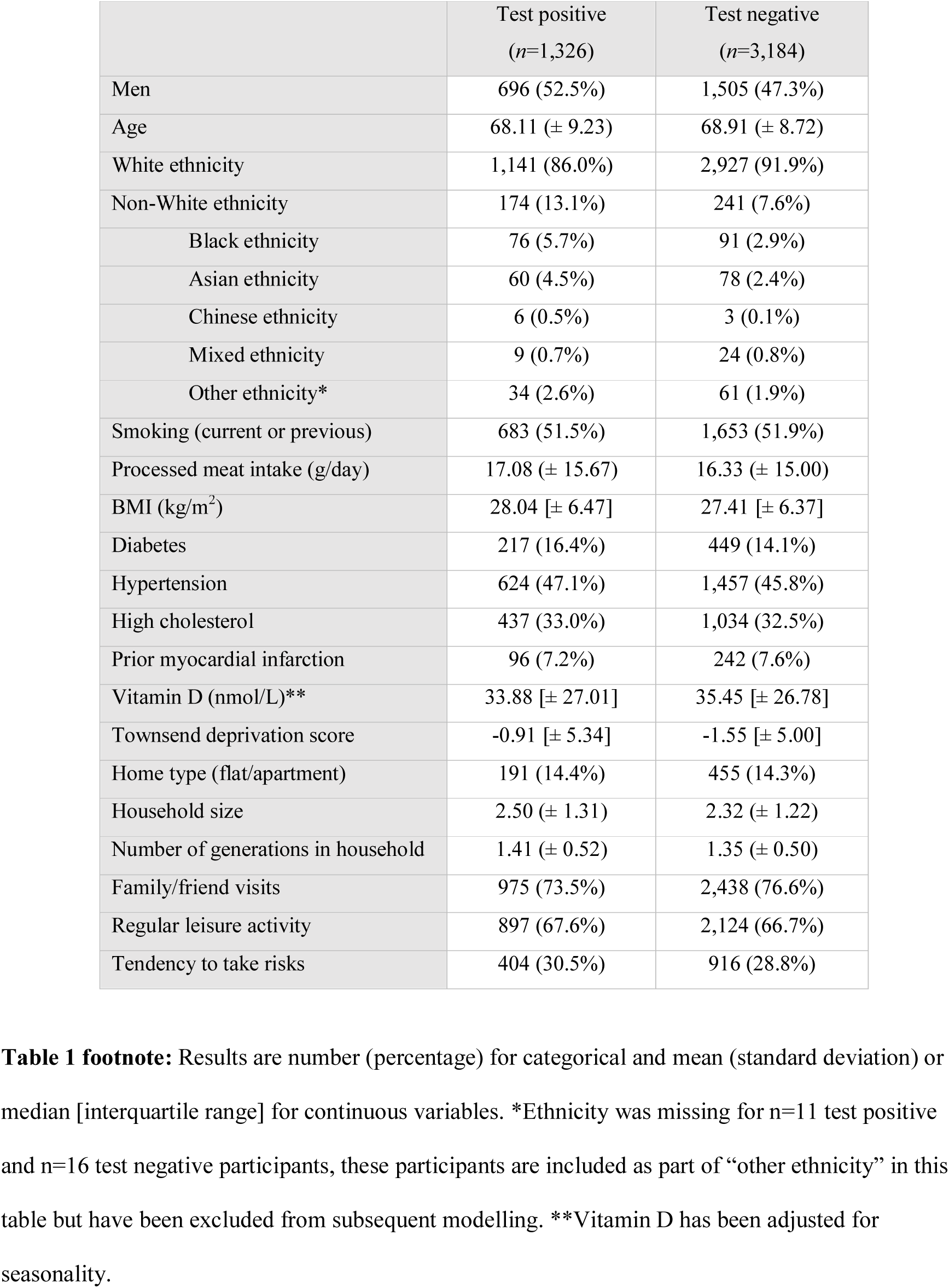
Baseline demographics by COVID-19 status.

footnote: Results are number (percentage) for categorical and mean (standard deviation) or median [interquartile range] for continuous variables.
|  | Test positive<br>(n=1,326) | Test negative<br>(n=3,184) |
| --- | --- | --- |
| Men | 696 (52.5%) | 1,505 (47.3%) |
| Age | 68.11 (± 9.23) | 68.91 (± 8.72) |
| White ethnicity | 1,141 (86.0%) | 2,927 (91.9%) |
| Non-White ethnicity | 174 (13.1%) | 241 (7.6%) |
| Black ethnicity | 76 (5.7%) | 91 (2.9%) |
| Asian ethnicity | 60 (4.5%) | 78 (2.4%) |
| Chinese ethnicity | 6 (0.5%) | 3 (0.1%) |
| Mixed ethnicity | 9 (0.7%) | 24 (0.8%) |
| Other ethnicity* | 34 (2.6%) | 61 (1.9%) |
| Smoking (current or previous) | 683 (51.5%) | 1,653 (51.9%) |
| Processed meat intake (g/day) | 17.08 (± 15.67) | 16.33 (± 15.00) |
| BMI (kg/m<sup>2</sup>) | 28.04 [± 6.47] | 27.41 [± 6.37] |
| Diabetes | 217 (16.4%) | 449 (14.1%) |
| Hypertension | 624 (47.1%) | 1,457 (45.8%) |
| High cholesterol | 437 (33.0%) | 1,034 (32.5%) |
| Prior myocardial infarction | 96 (7.2%) | 242 (7.6%) |
| Vitamin D (nmol/L)** | 33.88 [± 27.01] | 35.45 [± 26.78] |
| Townsend deprivation score | -0.91 [± 5.34] | -1.55 [± 5.00] |
| Home type (flat/apartment) | 191 (14.4%) | 455 (14.3%) |
| Household size | 2.50 (± 1.31) | 2.32 (± 1.22) |
| Number of generations in household | 1.41 (± 0.52) | 1.35 (± 0.50) |
| Family/friend visits | 975 (73.5%) | 2,438 (76.6%) |
| Regular leisure activity | 897 (67.6%) | 2,124 (66.7%) |
| Tendency to take risks | 404 (30.5%) | 916 (28.8%) |

#### Cardiometabolic factors and vitamin D

Men and non-White ethnicities had overall greater burden of cardiometabolic morbidities compared to women and White cohorts respectively (Figure 1E, Figure 1F). There were greater rates of smoking in White compared to Non-White ethnicities. Serum 25(OH)-vitamin D levels were, on average, higher in men and White ethnicities than women and non-White cohorts (Figure 1D).

#### Socio-demographic and behavioural factors

In comparison to the test negatives, those with a positive test had greater levels of social deprivation, were more likely to live in crowded households (Figure 1C). Non-Whites had higher levels of material deprivation by Townsend score compared to those of White ethnicity; levels of deprivation between men and women were similar (Supplementary Table 4). The frequency of family/friend visits and leisure activities outside the home were comparable between the test positive and test negative groups. There was greater tendency to risk taking behaviour in the test positive cohort, which was greater in men vs women and in non-White vs White ethnicities.

### Univariate associations of exposures with COVID-19 positive status

We tested the univariate association of all defined exposures with COVID-19 positive status within the tested cohort (Supplementary Table 6). Male sex, non-White ethnicity, higher BMI, greater material deprivation, and greater household overcrowding (household size, generations in household) were associated with increased odds of COVID-19 positive test. More frequent visits from family/friends were associated with lower risk of COVID-19 hospitalisation (given that a positive test implied hospital attendance), perhaps reflecting the role of social support in enabling individuals to remain at home when ill. There was a negative association between age and COVID-19 positivity, which may reflecting the narrow range and distribution of ages in the sample. Testing separately in men, non-White ethnicity, greater material deprivation, and higher BMI were the only statistically significant exposures. For women, additionally, lower 25(OH)-vitamin D status, greater household overcrowding (household size, generations in household), and greater risk taking behaviour were associated with COVID-19 positivity.

### Independent associations of specific exposures with COVID-19 status

#### Cardiometabolic factors

We undertook multivariate logistic regression models incorporating sex, age, ethnicity, smoking, BMI, diabetes, hypertension, high cholesterol, and prior MI (Table 2, Model 1). Male sex and non-White ethnicity were associated with greater odds of COVID-19 positive status with OR 1.28 (1.12, 1.46) and 1.78 (1.43, 2.20) respectively. Every 1kg/m^2^ of BMI was associated with 1.03 (1.01, 1.04) greater odds of COVID-19 positivity. There was a borderline negative association with age 0.99 [0.98, 1.00], which remained significant in sex-stratified analysis for women only. Non-White ethnicity and higher BMI were statistically significant associations for both men and women, with no evidence of attenuation (compared with the crude models).

**Table 2.**
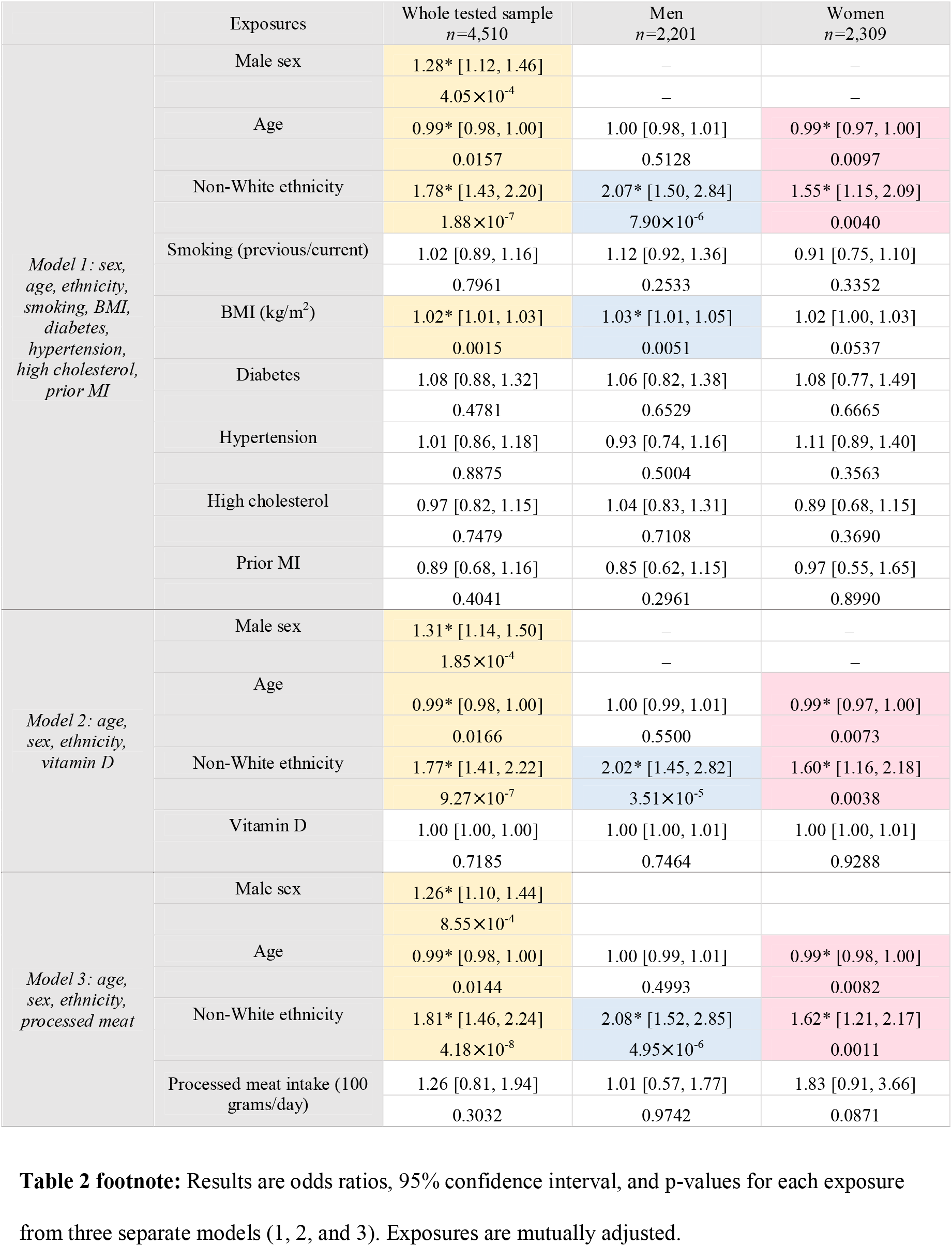
Multivariate logistic regression models testing the role of cardiometabolic factors (Model 1), vitamin D (Model 2), and poor diet (Model 3) in determining risk of COVID-19.

#### 25(OH)-vitamin D status and poor diet quality

In multivariate logistic regression models incorporating sex, age and ethnicity, there was no significant association between season-adjusted 25(OH)-vitamin D status and COVID-19 positivity (Table 2, Model 2). Similarly, in a separate model, adjustment for sex, age, and ethnicity demonstrated no statistically significant association between processed meat consumption and COVID-19 status (Table 2, Model 3). In both models, male sex was associated with higher odds of COVID-19 positive test and non-White ethnicity was consistently associated with COVID-19 positive test across men and women, with no evidence of attenuation.

#### Material deprivation

We tested the effect of material deprivation in multivariate models with mutual adjustment for sex, age, and ethnicity (Table 3, Model 4). There was a small, but statistically significant association between greater material deprivation and odds of COVID-19 positivity [OR 1.03 (1.01, 1.05)]. There remained strong and significant associations with male sex [OR 1.27 (1.11, 1.45)] and non-White ethnicity [OR 1.67 (1.34, 2.07)].

**Table 3.**
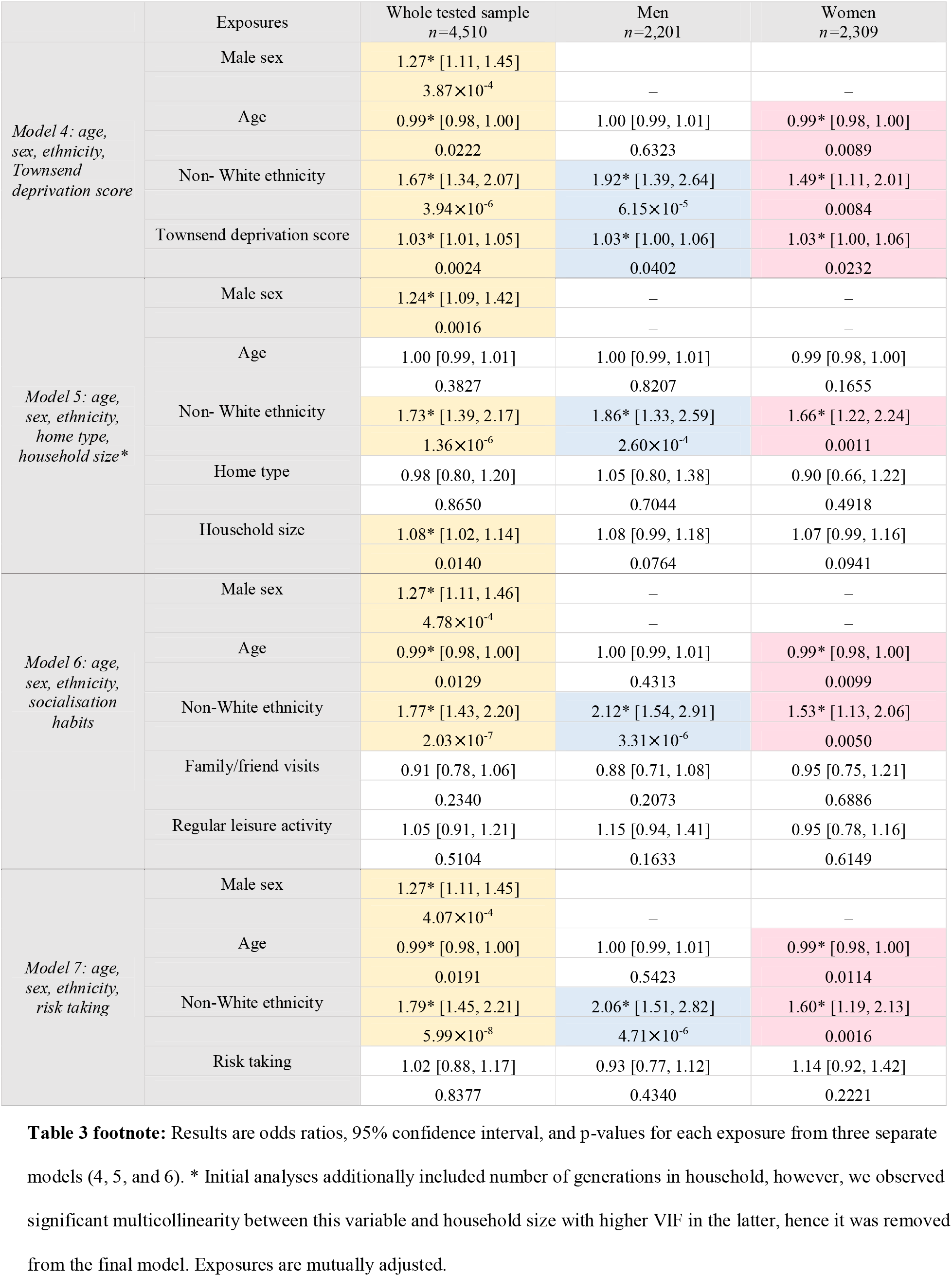
Multivariate logistic regression models testing the role of material deprivation (Model 4), housing conditions (Model 5), socialisation habits (Model 6), and attitude to risk (Model 7) in determining risk of COVID-19.

#### Housing conditions

We considered the effect of housing conditions in multivariate logistic regression models including sex, age, ethnicity, home type, and household size. In the whole sample, male sex, non-White ethnicity, and greater household size were associated with greater odds of COVID-19 positivity (Table 3, Model 5). Testing separately in men and women, non-White ethnicity was the only exposure which remained significantly associated with COVID-19 status. Attenuation of associations with household size is likely due to the small effect size and limited heterogeneity of the exposure in each of the sexes individually.

#### Socialisation habits and attitudes to risk

We undertook separate multivariate logistic regression models testing for associations between COVID-19 status, socialisation habits (Table 3, Model 6) and risk-taking attitude (Table 3, Model 7) whilst adjusting for age, sex, and ethnicity. Statistically significant associations were observed with male sex and non-White ethnicity which were not attenuated from crude models by adjustment for socialisation or risk-taking behaviour.

### Final model

We built a final multivariate logistic regression model, with covariates selected based on previous model permutations including sex, ethnicity, BMI, Townsend score, and household size (Supplementary Table 7). All covariates apart from age were statistically significant in this model. Male sex and non-White ethnicity were associated with greater odds of COVID-19 positivity: OR 1.23 (1.08, 1.41) and 1.59 (1.26, 1.99) respectively. Every 1kg/m^2^ increase in BMI was associated with 1.02 (1.01, 1.03) greater odds of COVID-19 positivity and for every additional person living in the same household the odds increased by 1.09 (1.03, 1.16).

## Discussion

### Main finding of this study

In 4,510 UKB participants tested for COVID-19 in a hospital setting (positive, *n* = 1,326), male sex, non-White ethnicity, higher BMI, and greater household size were associated with significantly greater odds of COVID-19 positive test. Despite variation in burden of cardiometabolic morbidities, 25(OH)-vitamin D levels, and material deprivation by sex and ethnicity, these factors were not significantly associated with COVID-19 positive status and did not explain the strong association with ethnicity. The pattern of tested associations did not vary between men and women.

### What is already known on this topic

There is mounting evidence for disproportionate adverse effects of COVID-19 in non-White ethnicities^2^. UK national audit data demonstrates that up to one-third of COVID-19 patients requiring intensive care are from BAME backgrounds, a rate which is far greater than their representation in the general population^17^. Similarly, an analysis of COVID-19 deaths amongst NHS staff, found that 64% of deaths were in individuals from BAME cohorts, markedly disproportionate to their 20% contribution to the NHS workforce^18^. The latest report from the Office of National Statistics (ONS) also demonstrates greater risk of COVID-19 mortality in BAME groups^19^; individuals of Black ethnicity appeared at highest risk with over 3.5 times greater risk of COVID-19 death compared to Caucasians, followed by Asian ethnicities^19^. Similarly, in the USA, there has been growing concern over the disproportionate impact of COVID-19 deaths amongst African Americans. In Chicago, 54.5% of all COVID-19 deaths are in Black people, whilst they constitute only 30% of the population^20^. These patterns are echoed across Europe, with Nordic countries reporting as much as ten times greater risk of COVID-19 in Somali populations^21^. We had previously documented this preponderance of cases amongst BAME individuals in our analysis of the initial UKB data release^16^; here we have confirmed the observation in the larger dataset, and importantly demonstrated a nonuniform impact across different BAME groups with highest rates amongst Black followed by Asian ethnicities.

The greater cardiometabolic burden in both BAME and male cohorts has been proposed as potentially important in driving adverse COVID-19 outcomes. In our sample, these cohorts had broadly poorer cardiometabolic profile than their female and White counterparts. However, in multivariate logistic regression models including multiple cardiometabolic factors statistically significant associations with COVID-19 status were observed with male sex, non-White ethnicity, and higher BMI only. This suggests that the greater cardiometabolic burden in non-White ethnicities does not account for the adverse COVID-19 outcomes in this group.

Consistent with our findings, UK and USA statistics highlight obesity as a marker of poor COVID-19 outcomes, such as requirement for intensive care^22^. There are increasing suggestions of a possible pathophysiological link between adiposity and COVID-19 severity. Wide expression of ACE2 receptors within adipose tissue is thought to promote binding and cellular entry of severe acute respiratory syndrome coronavirus 2 (SARS-CoV-2)^23^. It has been suggested that adipose tissue may act as a “viral reservoir” for SARS-CoV-2, perhaps contributing to a more prolonged and severe illness^23^. In addition, adipose tissue is a known source of inflammatory cytokines, such as Interleukin 6^24^. This is hypothesised to be linked to the association of adiposity with greater likelihood of cytokine storms and the consequent risk of severe respiratory complications in the context of COVID-19. Indeed, studies have demonstrated association of higher Interleukin 6 levels with respiratory failure and requirement for mechanical ventilation in COVID-19 patients^25^. Greater adiposity, as well as non-White ethnicity, is associated with lower 25(OH)-vitamin D status. Although the active 1,25(OH)_2_-vitamin D form has immune system functions^26^, evidence linking low 25(OH-vitamin D (the circulating storage form, and poorly correlated with 1,25(OH)_2_-vitamin D) with COVID-19 disease have been contradictory^27^. In our study, our finding of no independent associations between 25(OH)-vitamin D status and COVID-19 disease suggest that the association is confounded by ethnicity and BMI. Interestingly, the BMI association was retained in multivariate models, suggesting a possible independent role for adiposity, which clearly deserves further investigation.

Socio-economic deprivation is associated with poorer global health outcomes^28^ with specific implications for infectious disease due to its association with overcrowded living conditions. It has been suggested that ethnic differences in COVID-19 severity may relate to clustering of material deprivation with BAME status^29^. In the UKB material deprivation is reported using the Townsend score, which is based on four factors-employment, car ownership, home ownership, and household overcrowding. Consistent with national reports, we found higher material deprivation in non-White participants. In multivariate models including age, sex, ethnicity, and Townsend score, there were significantly greater odds of COVID-19 with greater material deprivation whilst the association with ethnicity appeared strong and significant. Testing separately for the effect of household overcrowding, this exposure appeared significant independent of sex, ethnicity, age, and home type. This suggests that in modern western societies, it is not global economic deprivation, but specific aspects relating to household overcrowding that have relevance to COVID-19. Consistent with these observations, a survey of COVID-19 cases from New York reports the highest number of cases occurring in areas with the largest average household size^30^. Furthermore, analysis of UK cases by the ONS also demonstrates that material deprivation does not adequately explain the ethnic disparities in COVID-19 outcomes^19^.

Behavioural factors, in particular attitudes that may compromise adherence to lockdown measures, have been proposed as potentially important in determining risk of exposure to SARS-CoV-2. There have been suggestions that the higher rates of COVID-19 in men may relate to their greater tendency to take risks than women^31^ with a recent UK survey identifying young men as the most likely demographic to break lockdown measures^32^. Cultural differences in socialisation habits and more frequent contact with friends and extended family are also potentially relevant factors. In our analysis, we did not find these factors to be significantly important in conferring COVID-19 positive status with the increased odds associated with male sex and non-White ethnicity occurring independent of these factors.

### What this study adds

This study is consistent with growing reports of higher risk of severe COVID-19 in men and non-White ethnicities, in particular Black populations. We identified higher BMI, greater material deprivation, and household overcrowding as independent risk factors for COVID-19. In addition, we demonstrated that the ethnic and sex differential pattern of COVID-19 is not adequately explained by greater cardiometabolic disease burden, lower vitamin D levels, greater social deprivation, or behavioural factors. Thus, alternative biological pathways and different genetic susceptibilities warrant consideration.

### Limitations of this study

Aggregating all BAME populations into one cohort may overlook important differences between non-Caucasian ethnicities. Further study in samples with greater ethnic diversity is needed. Given the observational nature of the study, we cannot discern causal relationships, and although we controlled for a wide range of covariates, the possibility of residual confounding should be considered. Furthermore, the current dataset does not allow assessment of specific COVID-19 health outcomes.

## Conclusion

There is greater risk of severe COVID-19 amongst men and non-White ethnicities. The augmented risk in BAME populations is non-uniform and disproportionately affects Black and Asian ethnicities. The sex and ethnicity differential pattern of COVID-19 is not adequately explained by variations in cardiometabolic factors, 25(OH)-vitamin D levels, or socio-economic factors. Thus, investigation of alternative biological pathways and genetic susceptibilities is warranted.

## Data Availability

Data in this work is from the UK Biobank. This dataset is available to all bone fide researchers on completion of a formal access application, which can be found on the UK Biobank website: https://www.ukbiobank.ac.uk.

## Acknowledgements

This study was undertaken using the UK Biobank resource, Access Application 2964.

